# Glycemic Fluctuations, Fatigue, and Sleep Disturbances in Type 2 Diabetes During Ramadan Fasting: A Cross-Sectional Study

**DOI:** 10.1101/2024.10.08.24315087

**Authors:** Satwika Arya Pratama, Rudy Kurniawan, Hsiao-Yean Chiu, Hsuan-Ju Kuo, Emmanuel Ekpor, Safiruddin Al-Baqi, Faizul Hasan, Debby Syahru Romadlon

## Abstract

**Background:** This study aimed to assess the prevalence of glycemic fluctuations, fatigue, and sleep disturbances during Ramadan, and to identify factors associated with hypoglycemia and hyperglycemia events in this period.

**Methods:** A cross-sectional study of 88 individuals with type 2 diabetes during Ramadan fasting (March-April 2024) was conducted. HbA1c levels before Ramadan were obtained from medical records. Participants monitored blood glucose twice daily (during the day and two hours after breaking fast). Blood glucose under 70 mg/dl was considered hypoglycemia, and over 200 mg/dl was hyperglycemia. Fatigue and sleep quality were assessed using the Indonesian Multidimensional Fatigue Inventory-20 (IMFI-20) and the Pittsburgh Sleep Quality Index (PSQI).

**Results:** A total of 88 patients with type 2 diabetes (mean age, 52.7 years) participated, predominantly female (68.2%) and married (63.6%). The study found a prevalence of 21.6% for hypoglycemia and 30.6% for hyperglycemia. Additionally, 30.7% of participants experienced fatigue, and 40.9% reported poor sleep quality. HbA1c levels before Ramadan and fatigue were significantly associated with both hypoglycemia and hyperglycemia (p < 0.05). Sleep quality was also significantly associated with hyperglycemia events (p < 0.05).

**Conclusion:** The study found that hypoglycemia and hyperglycemia are common in individuals with type 2 diabetes during Ramadan. Fatigue and poor sleep quality were also prevalent.

Significant factors linked to both glycemic events included pre-Ramadan HbA1c levels and fatigue, while sleep quality was associated with hyperglycemia. These findings emphasize the need for targeted interventions and personalized management to reduce glycemic fluctuations and improve well-being during Ramadan fasting.

## 1 INTRODUCTION

Type 2 diabetes mellitus is a prevalent chronic metabolic disorder characterized by insulin resistance and relative insulin deficiency, leading to hyperglycemia [1]. It is associated with various complications, including cardiovascular diseases, neuropathy, retinopathy, and nephropathy, which significantly impact the quality of life and overall health outcomes of affected individuals [2]. The management of type 2 diabetes typically involves lifestyle modifications, pharmacotherapy, and continuous monitoring of blood glucose levels to mitigate the risks of complications. Among the lifestyle modifications, dietary habits play a crucial role in maintaining glycemic control [3]. Ramadan, the holy month of fasting observed by Muslims worldwide, poses unique challenges for individuals with type 2 diabetes due to the significant alterations in meal patterns and fasting durations.

Fasting during Ramadan involves abstaining from food and drink from dawn until sunset, which can lead to substantial changes in blood glucose levels [4]. These fluctuations in glycemia can exacerbate symptoms of fatigue and disturb sleep patterns in individuals with type 2 diabetes [5]. Previous studies have shown that Ramadan fasting can result in both hyperglycemia and hypoglycemia, posing risks for patients with diabetes [6]. Moreover, the alterations in circadian rhythms and sleep-wake cycles during Ramadan can further complicate the management of type 2 diabetes, leading to increased fatigue and sleep disturbances [7]. Therefore, it is imperative to understand the impact of Ramadan fasting on glycemic control and associated symptoms in this population.

Fatigue is a common and debilitating symptom experienced by individuals with type 2 diabetes, often exacerbated by poor glycemic control [8-10]. It can significantly impair daily functioning and quality of life, making it a critical aspect of diabetes management. The relationship between glycemic fluctuations and fatigue is complex, involving various physiological and psychological mechanisms. Hyperglycemia can lead to osmotic diuresis and dehydration, resulting in fatigue, while hypoglycemia can cause neuroglycopenic symptoms such as weakness and lethargy [11]. Additionally, sleep disturbances, which are prevalent among individuals with type 2 diabetes, can further aggravate fatigue and disrupt overall metabolic control [12]. This study aimed to investigate the prevalence of glycemic fluctuations, fatigue, and sleep disturbances during Ramadan and to examine the factors associated with hypoglycemia and hyperglycemia events during this period

Understanding the interplay between glycemic control, fatigue, and sleep disturbances during Ramadan is crucial for developing tailored management strategies for individuals with type 2 diabetes who choose to fast. While several studies have investigated the effects of Ramadan fasting on glycemic control, there is a paucity of research focusing on the concurrent impact on fatigue and sleep disturbances. By conducting a cross-sectional study, this research seeks to fill this gap in the literature and provide insights into the challenges faced by patients with diabetes during Ramadan. The findings of this study could inform healthcare providers in offering comprehensive guidance and support to patients with type 2 diabetes, ensuring safer fasting practices and improved overall well-being during Ramadan [13].

## 2 METHODS

### 2.1 Study design and setting

This study was a cross-sectional investigation utilizing convenience sampling, conducted at a diabetes management center in Indonesia during the Ramadan fasting period in March and April 2024. The research received approval from the Joint Institutional Review Board of the Ethical Committee of Medical Research at the Faculty of Dentistry, University Jember (No.2458/UN25.8/KEPK/DL/2024). Written informed consent was obtained from all participants who agreed to take part in the survey. Additionally, the study adhered to the Strengthening the Reporting of Observational Studies in Epidemiology (STROBE) guidelines [14] (S1 Table).

### 2.2 Study populations

Participants were recruited from a diabetes management center in Indonesia. We utilized elements for risk calculation and the suggested risk score for individuals with diabetes mellitus who wish to fast during Ramadan [15]. Only those with a low to moderate risk score, and who chose to fast during Ramadan, were included in the study. Additionally, participants were required to be between 17 (the legal age for providing informed consent in Indonesia) and 65 years old, and to own a mobile phone. Individuals who could not read or write Indonesian, or who had been diagnosed with cognitive impairment, a psychiatric disorder, or cancer prior to the study, were excluded.

### 2.3 Outcomes

#### 2.3.1 Blood glucose levels

Glucometers were used by the participants to monitor their own blood glucose levels twice daily (during the day and two hours after break fasting) during Ramadan [15]. The participants were invited to join a WhatsApp group to report their blood glucose levels each day during Ramadan. Blood glucose levels below 70 mg/dl were classified as hypoglycemia, and those above 200 mg/dl were considered hyperglycemia. Participants who experienced hypoglycemia were asked to break their fast during the period of fasting. Furthermore, HbA1c levels before Ramadan were collected from the medical records of the participants.

#### 2.3.2 Fatigue

The fatigue levels of participants during Ramadan were measured using the Indonesian version of the Multidimensional Fatigue Inventory-20 (IMFI-20) [9]. The IMFI-20 consists of 20 items across 4 subscales: general and physical fatigue, reduced motivation, reduced activity, and mental fatigue. Each subscale includes 4 items rated on a 5-point Likert scale, ranging from 1 (strongly agree) to 5 (strongly disagree). We reverse-scored 10 positively worded items (items 2, 5, 9, 10, 13, 14, 16, 17, 18, and 19). The score for each subscale (ranging from 4 to 20 points) was the sum of its item scores, and the total fatigue score (ranging from 20 to 100 points) was the sum of the subscale scores, with higher scores indicating greater fatigue. The IMFI-20 demonstrated high validity and reliability (Cronbach’s α = 0.92).

### 2.3.3 Sleep Quality

Sleep quality among individuals with type 2 diabetes during Ramadan fasting was assessed using the Pittsburgh Sleep Quality Index (PSQI). The PSQI measures self-reported sleep quality and disturbances over the past month. It consists of 19 items across 7 dimensions: (1) subjective sleep quality, (2) sleep latency, (3) sleep duration, (4) sleep efficiency, (5) sleep disturbances, (6) use of sleeping medication, and (7) daytime dysfunction. Items are rated on a 4-point Likert scale ranging from 0 to 3, with an overall score range of 0 to 20. A score below 5 indicates good sleep quality [16]. The Indonesian version of the PSQI has demonstrated high validity and reliability (Cronbach’s α = 0.72) [17].

#### 2.3.4 Demographic and disease characteristics and self-reported fatigue and sleep quality during Ramadan fasting

Predesigned information sheets were employed to gather data on demographic and disease characteristics, including age, sex, education level, marital status, income level, and current diabetes treatment. Additionally, we included questions regarding the participants’ experiences of fatigue and sleep quality, as well as the duration of these experiences during Ramadan fasting in a healthcare setting (S2 Table).

### 2.4 Data collection

Prior to Ramadan fasting, we invited potential participants at the diabetes management center to undergo a fasting risk assessment [15] and inquired about their intention to fast.

Informed consent was obtained from those who agreed to participate. We also collected the WhatsApp numbers of all participants and created a WhatsApp group for reporting blood glucose levels during Ramadan. In the week following Ramadan, we invited participants back to the diabetes management center to complete the questionnaires and demographic form again. All participants who completed the study received a small gift.

### 2.5 Statistical analysis

All analyses were conducted using SPSS version 24.0 (IBM, Armonk, NY, USA), with a P value of <0.05 considered statistically significant. Demographic and disease characteristics were presented as means and standard deviations for continuous variables and as numbers and percentages for categorical variables. After consulting with the physicians, we assessed four factors associated with hypoglycemia and hyperglycemia events during Ramadan fasting, which included type of medications, comorbidity, HbA1c levels before Ramadan, fatigue, and sleep quality. The Chi-square test was employed to evaluate the associations between these factors and hypoglycemia and hyperglycemia events. An independent t-test was used to examine the relationship between the total IMFI-20 score and its domains with hypoglycemia and hyperglycemia events during Ramadan.

## 3 RESULTS

### 3.1 General characteristics of the participants in the health care setting

A total of 88 patients with type 2 diabetes (mean age of 52.7 years) participated in this study. The majority were female (68.2%) and married (63.6%). Approximately 89.8% of the participants were solely on oral hypoglycemic agents (OHA), with 58% having comorbid conditions. About 58% of the participants fasted for all 30 days of Ramadan, while 22.7% fasted for 15-29 days. The mean of HbA1c level before Ramadan was 6.7 mg/dl. Detailed participant characteristics are presented in Table 1.

**Table 1.** Participant characteristics and diseases (N = 88)

| **Variables** | **Total** |
| --- | --- |
| Age | 52.7 (5.5) |
| Female | 60 (68.2) |
| Diabetes duration (years) | 5.7 (2.5) |
| Junior high school and above | 52 (59.1) |
| Married | 56 (63.6) |
| Having comorbidity | 51 (58.0) |
| Medications |  |
| OHA alone | 79 (89.8) |
| OHA combined with Insulin | 9 (10.2) |
| Duration of fasting during Ramadan |  |
| 1-14 days | 17 (19.3) |
| 15-29 days | 20 (22.7) |
| 30 days | 51 (58.0) |
| HbA1c (%) before Ramadan | 6.7 (0.5) |

Continuous variables are presented in mean and standard deviation.
Categorical variables are presented in number (%)

### 3.2 Factor associated with glycemic fluctuations during Ramadan

Table 2 presents that the prevalence of hypoglycemia and hyperglycemia in type 2 diabetes during Ramadan was 21.6% and 30.6%, respectively. Additionally, 30.7% of participants experienced fatigue during Ramadan fasting, and 40.9% reported poor sleep quality during this period. The HbA1c levels of participants before Ramadan were significantly associated with both hypoglycemia (p = 0.03) and hyperglycemia cases (p < 0.001). Participants with higher HbA1c before Ramadan scores were more likely to experience hyperglycemia during Ramadan. Fatigue symptoms were significantly linked to both hypoglycemia and hyperglycemia events during Ramadan (p < 0.05), with most participants who reported fatigue also having abnormal blood glucose levels.

**Table 2.**
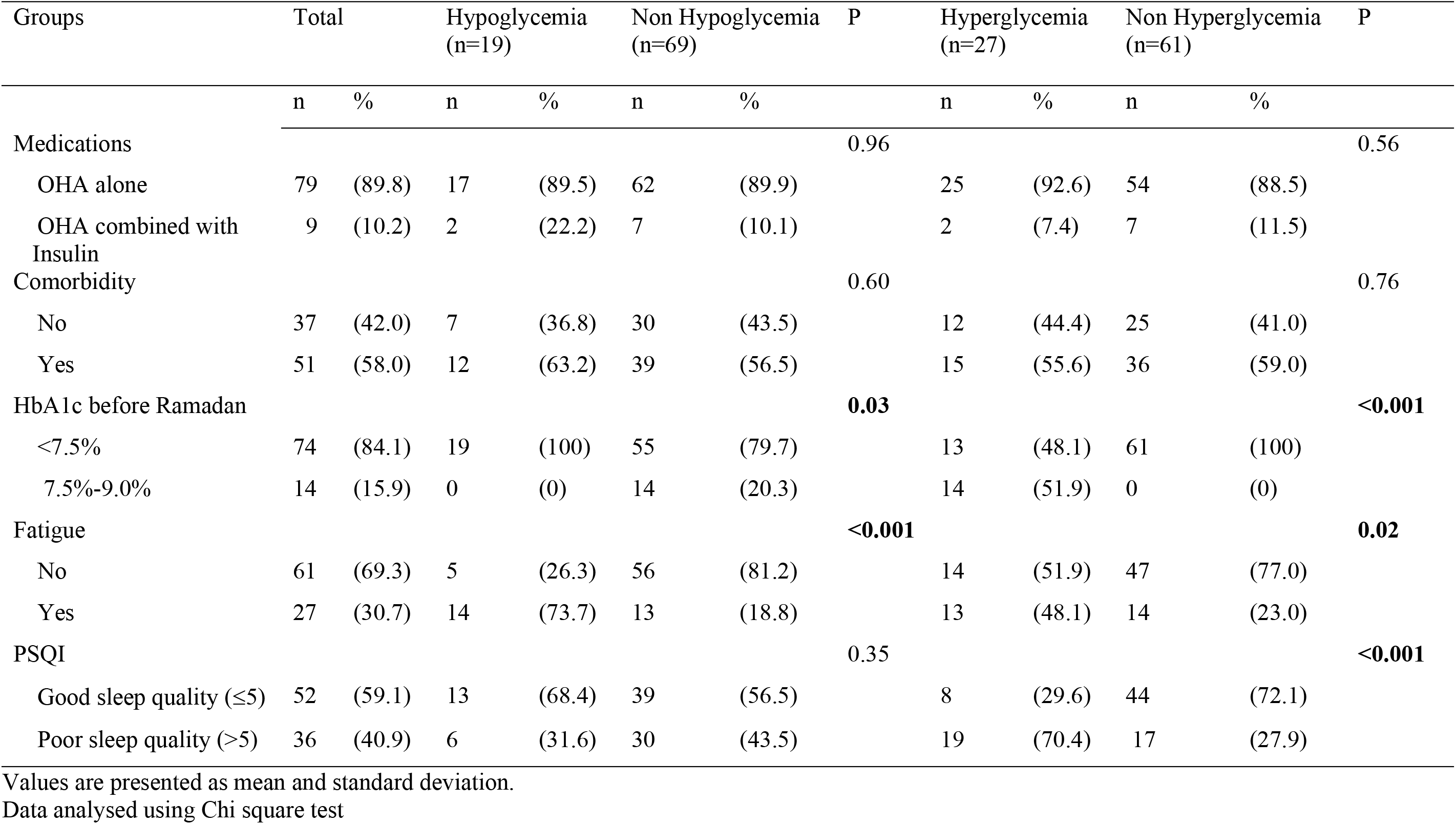
Relationship between glycemic fluctuations with factors during Ramadan fasting.

Furthermore, sleep quality was significantly associated with hyperglycemia events (p < 0.0001). However, comorbidity and the type of medications did not show a significant association with hypoglycemia and hyperglycemia events during Ramadan (p > 0.05, Table 2). Table 3 indicates that both hypoglycemia and hyperglycemia were significantly associated with the total score of the IMFI-20 (p < 0.05). Hypoglycemia was linked to all three domains of the IMFI-20, including general/physical fatigue, mental fatigue, and reduced activity (p < 0.05), whereas hyperglycemia was only significantly associated with general/physical fatigue during Ramadan, as detailed in Table 3.

**Table 3.** IMFI-20 and glycemic fluctuations during Ramadan.

| Groups | n | IMFI-20 | General/physical fatigue | Mental fatigue | Reduced Activity | Reduced Motivation |
| --- | --- | --- | --- | --- | --- | --- |
| Hypoglycemia |  |  |  |  |  |  |
| No | 69 | 53.3 ± 12.2* | 24.4 ± 7.5 | 10.1 ± 1.8* | 8.9 ± 2.5* | 9.6 ± 4.3 |
| Yes | 19 | 60.0 ± 11.5* | 27.2 ± 7.8 | 11.1 ± 2.2* | 10.7 ± 2.4* | 11.2 ± 4.6 |
| Hyperglycemia |  |  |  |  |  |  |
| No | 61 | 52.8 ± 10.9* | 23.8 ± 6.6* | 10.2 ± 2.0 | 9.3 ± 2.7 | 9.5 ± 3.7 |
| Yes | 27 | 58.9 ± 14.2* | 28.3 ± 8.7* | 10.5 ± 1.6 | 9.4 ± 2.3 | 11.0 ± 5.4 |

Data analysed using independent t test

### 3.3 Fatigue and sleep quality among type 2 diabetes during Ramadan in health care settings

Concerning fatigue and sleep quality among individuals with type 2 diabetes during Ramadan (as shown in Tables 4 and 5), 46.7% of participants who experienced fatigue and 75% of those with poor sleep quality during Ramadan fasting rarely or never discussed these issues with their physicians. Additionally, participants with fatigue (81.5%) and poor sleep quality (86.1%) were rarely or never treated by their physicians.

**Table 4.** Healthcare service usage for addressing fatigue among people with type 2 diabetes during Ramadan.

| Questions | Total |  | Fatigued (n= 27) |  | Non-Fatigued (n=61) |  |
| --- | --- | --- | --- | --- | --- | --- |
|  | n | % | n | % | n | % |
| How long have you felt fatigue during Ramadan? (day), mean (SD) | 1.8 | 2.9 | 5.9 | 2.0 | 00.0 | 0.0 |
| Have you ever discussed “the feeling of tired” or “fatigue” with your physician during Ramadan? |  |  |  |  |  |  |
| Never | 69 | (78.4) | 8 | (9.6) | 61 | (100) |
| Rarely | 10 | (11.4) | 10 | (37.1) | 0 | (0) |
| Sometimes | 5 | (5.7) | 5 | (18.5) | 0 | (0) |
| Often | 4 | (4.5) | 4 | (14.8) | 0 | (0) |
| Always | 0 | (0) | 0 | (0) | 0 | (0) |
| Whether your fatigue level has been measured by your physician during Ramadan? |  |  |  |  |  |  |
| Never | 75 | (85.2) | 14 | (51.9) | 61 | (100) |
| Rarely | 8 | (9.1) | 8 | (29.6) | 0 | (0) |
| Sometimes | 5 | (5.7) | 5 | (18.5) | 0 | (0) |
| Often | 0 | (0) | 0 | (0) | 0 | (0) |
| Always | 0 | (0) | 0 | (0) | 0 | (0) |
| Has your fatigue been treated by physician during Ramadan? |  |  |  |  |  |  |
| Never | 73 | (82.9) | 12 | (44.4) | 61 | (100) |
| Rarely | 10 | (11.4) | 10 | (37.1) | 0 | (0) |
| Sometimes | 5 | (5.7) | 5 | (18.5) | 0 | (0) |
| Often | 0 | (0) | 0 | (0) | 0 | (0) |
| Always | 0 | (0) | 0 | (0) | 0 | (0) |

**Table 5.**
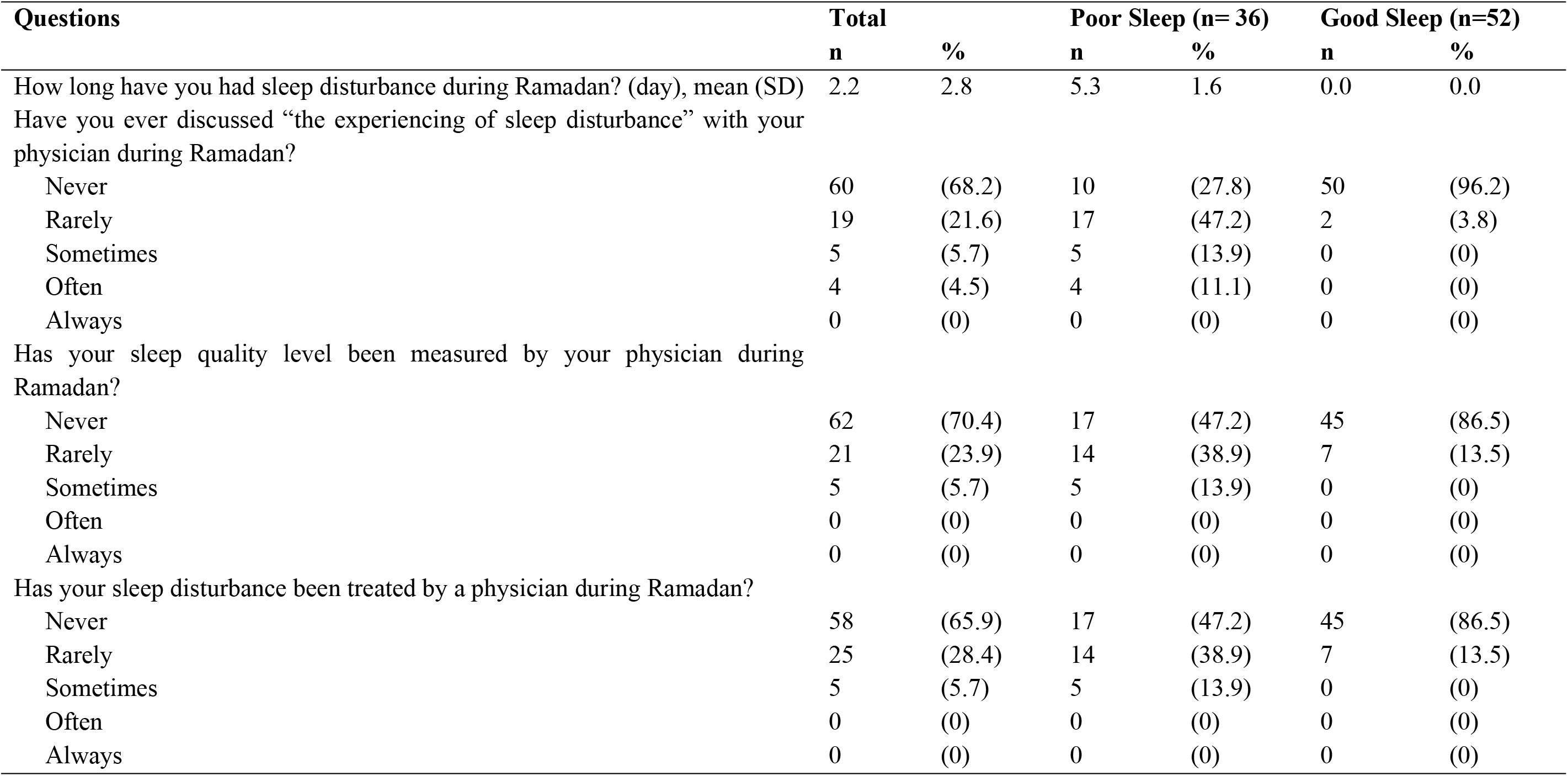
Healthcare service usage for addressing sleep quality among people with type 2 diabetes during Ramadan.

| Questions | Total |  | Poor Sleep (n= 36) |  | Good Sleep (n=52) |  |
| --- | --- | --- | --- | --- | --- | --- |
|  | n | % | n | % | n | % |
| How long have you had sleep disturbance during Ramadan? (day), mean (SD) | 2.2 | 2.8 | 5.3 | 1.6 | 0.0 | 0.0 |
| Have you ever discussed “the experiencing of sleep disturbance” with your physician during Ramadan? |  |  |  |  |  |  |
| Never | 60 | (68.2) | 10 | (27.8) | 50 | (96.2) |
| Rarely | 19 | (21.6) | 17 | (47.2) | 2 | (3.8) |
| Sometimes | 5 | (5.7) | 5 | (13.9) | 0 | (0) |
| Often | 4 | (4.5) | 4 | (11.1) | 0 | (0) |
| Always | 0 | (0) | 0 | (0) | 0 | (0) |
| Has your sleep quality level been measured by your physician during Ramadan? |  |  |  |  |  |  |
| Never | 62 | (70.4) | 17 | (47.2) | 45 | (86.5) |
| Rarely | 21 | (23.9) | 14 | (38.9) | 7 | (13.5) |
| Sometimes | 5 | (5.7) | 5 | (13.9) | 0 | (0) |
| Often | 0 | (0) | 0 | (0) | 0 | (0) |
| Always | 0 | (0) | 0 | (0) | 0 | (0) |
| Has your sleep disturbance been treated by a physician during Ramadan? |  |  |  |  |  |  |
| Never | 58 | (65.9) | 17 | (47.2) | 45 | (86.5) |
| Rarely | 25 | (28.4) | 14 | (38.9) | 7 | (13.5) |
| Sometimes | 5 | (5.7) | 5 | (13.9) | 0 | (0) |
| Often | 0 | (0) | 0 | (0) | 0 | (0) |
| Always | 0 | (0) | 0 | (0) | 0 | (0) |

## 4 DISCUSSION

This recent study found that hypoglycemia and hyperglycemia are prevalent among individuals with type 2 diabetes during Ramadan fasting. Additionally, some of the participants experienced fatigue and poor sleep quality during this period. The HbA1c level before Ramadan and fatigue were significantly associated with the occurrence of both hypoglycemia and hyperglycemia during Ramadan. Poor sleep quality was also a significant factor associated with hyperglycemia during this time.

The findings of our study indicate that hyperglycemia was more prevalent than hypoglycemia during Ramadan. These results highlight the dual challenge faced by individuals with glucose regulation issues during Ramadan fasting [18]. Previous studies have documented similar challenges in patients with type 1 diabetes [19], but our study adds to the growing body of evidence by quantifying the extent of these issues in type 2 diabetes. The high prevalence underscores the need for tailored medical advice and monitoring for individuals with prediabetes or diabetes during Ramadan.

Our analysis revealed that fatigue and poor sleep quality are prevalent among the fasting individuals. These factors are critical as they not only impact daily functioning but also appear to be linked to glucose regulation. The association between fatigue and both hypoglycemia and hyperglycemia suggests that physical and mental exhaustion could influence metabolic control during Ramadan. This is consistent with the understanding that stress and lack of rest can exacerbate glucose fluctuations [20], emphasizing the importance of managing overall health and well-being during fasting.

Importantly, our study identified HbA1c levels prior to Ramadan as significant factors associated with hypoglycemia and hyperglycemia events during fasting. This finding suggests that individuals with higher baseline HbA1c levels are more susceptible to glucose fluctuations, which could be attributed to their overall poorer glycemic control. These insights are crucial for healthcare providers in developing personalized strategies to minimize the risks of hypoglycemia and hyperglycemia for fasting individuals.

Sleep quality also emerged as a significant factor associated with hyperglycemia events during Ramadan. Poor sleep quality can lead to hormonal imbalances that affect insulin sensitivity and glucose metabolism [21], thereby increasing the risk of hyperglycemia. This relationship underscores the importance of adequate and quality sleep in managing glucose levels. Interventions aimed at improving sleep hygiene could, therefore, be beneficial for individuals fasting during Ramadan. Future research should explore targeted interventions to enhance sleep quality and reduce fatigue as part of comprehensive diabetes management during fasting periods.

### 4.1 Limitation and strengths

This study has several limitations. Firstly, the participant pool was limited to individuals residing in urban areas, potentially restricting the broader applicability of the findings. Secondly, the cross-sectional design of this study makes it challenging to establish causality between exposure and outcomes. Despite these limitations, this research is pioneering in investigating the relationship between fatigue, sleep quality, and glycemic fluctuations among individuals with type 2 diabetes during Ramadan fasting.

## 5 CONCLUSION

This study highlights the significant prevalence of hypoglycemia and hyperglycemia among individuals with type 2 diabetes during Ramadan fasting. Additionally, fatigue and poor sleep quality were prevalent among people with type 2 diabetes during Ramadan. Importantly, HbA1c levels before Ramadan and fatigue were found to be significant factors associated with both hypoglycemia and hyperglycemia. Furthermore, sleep quality was identified as a significant factor contributing to hyperglycemia events. These findings underscore the need for targeted interventions and personalized management strategies to mitigate glycemic fluctuations and improve overall well-being for individuals with type 2 diabetes during Ramadan fasting. Future research should focus on developing comprehensive care plans that address these critical factors.

## FUNDING

The authors received no specific funding for this work.

## ACKNOWLEDGMENTS

None.

## CONFLICTS OF INTEREST

The authors declare no conflicts of interest.

## AUTHOR CONTRIBUTIONS

SAP, RK, SS, and DSR designed the study concept; SAP, RK, SAB, and DSR extracted the data; SAP, RK, and DSR performed the statistical analyses; SAP, RK, HYC, HJK, EE, SAB, FH, SS, and DSR assessed the study quality; SAP, RK, SS, and DSR wrote the original manuscript; SAP, RK, SS, and DSR revised the final manuscript; all authors have read and approved this final manuscript.

## DATA AVAILABILITY STATEMENT

Specific data files used in the analysis are available from the corresponding author upon reasonable request.

## Notes

### Competing Interest Statement

The authors have declared no competing interest.

### Funding Statement

The author(s) received no specific funding for this work.

### Author Declarations

The research received approval from the Joint Institutional Review Board of the Ethical Committee of Medical Research at the Faculty of Dentistry, University Jember (No.2458/UN25.8/KEPK/DL/2024).

## REFERENCES

1. Galicia-Garcia, U., et al., Pathophysiology of type 2 diabetes mellitus. International journal of molecular sciences, 2020. 21(17): p. 6275. DOI: 10.3390/ijms21176275

2. Association, A.D., 2. Classification and diagnosis of diabetes: standards of medical care in diabetes—2020. Diabetes care, 2020. 43(Supplement_1): p. S14–S31. DOI: 10.2337/dc21-S002

3. Dyson, P., et al., Diabetes UK evidence-based nutrition guidelines for the prevention and management of diabetes. Diabetic medicine, 2018. 35(5): p. 541–547. DOI: 10.1111/dme.13603

4. Ahmed, S.H., et al., Ramadan and diabetes: a narrative review and practice update. Diabetes Therapy, 2020. 11: p. 2477–2520. DOI: 10.1007/s13300-020-00886-y

5. Salti, I., et al., A population-based study of diabetes and its characteristics during the fasting month of Ramadan in 13 countries: results of the epidemiology of diabetes and Ramadan 1422/2001 (EPIDIAR) study. Diabetes care, 2004. 27(10): p. 2306–2311. DOI: 10.2337/diacare.27.10.2306

6. Al-Arouj, M., et al., Recommendations for management of diabetes during Ramadan: update 2010. Diabetes care, 2010. 33(8): p. 1895. DOI: 10.2337/dc10-0896

7. Hassanein, M., et al., Diabetes and Ramadan: practical guidelines. Diabetes research and clinical practice, 2017. 126: p. 303–316. DOI: 10.1016/j.diabres.2021.109185

8. Romadlon, D.S., et al., Prevalence and risk factors of fatigue in type 1 and type 2 diabetes: A systematic review and meta-analysis. Journal of Nursing Scholarship, 2021. DOI: 10.1111/jnu.12763

9. Romadlon, D.S., et al., Fatigue following type 2 diabetes: Psychometric testing of the Indonesian version of the multidimensional fatigue Inventory-20 and unmet fatigue-related needs. Plos one, 2022. 17(11): p. e0278165. DOI: 10.1371/journal.pone.0278165

10. Fritschi, C. and L. Quinn, Fatigue in patients with diabetes: a review. Journal of psychosomatic research, 2010. 69(1): p. 33–41. DOI: 10.1016/j.jpsychores.2010.01.021

11. Barendse, S., et al., The impact of hypoglycaemia on quality of life and related patient-reported outcomes in Type 2 diabetes: a narrative review. Diabetic medicine, 2012. 29(3): p. 293–302. DOI: 10.1111/j.1464-5491.2011.03416.x

12. Luyster, F.S. and J. Dunbar-Jacob, Sleep quality and quality of life in adults with type 2 diabetes. The diabetes educator, 2011. 37(3): p. 347–355. DOI: 10.1177/0145721711400663

13. Benaji, B., et al., Diabetes and Ramadan: review of the literature. Diabetes research and clinical practice, 2006. 73(2): p. 117–125. DOI: 10.1016/j.diabres.2005.10.028

14. Von Elm, E., et al., The Strengthening the Reporting of Observational Studies in Epidemiology (STROBE) Statement: guidelines for reporting observational studies. International journal of surgery, 2014. 12(12): p. 1495–1499. DOI: 10.1016/j.jclinepi.2007.11.008

15. Hassanein, M., et al., Diabetes and Ramadan: practical guidelines 2021. Diabetes research and clinical practice, 2022. 185: p. 109185. DOI: 10.1016/j.diabres.2021.109185

16. Buysse, D.J., et al., The Pittsburgh Sleep Quality Index: a new instrument for psychiatric practice and research. Psychiatry research, 1989. 28(2): p. 193–213. DOI: 10.1016/0165-1781(89)90047-4

17. Setyowati, A. and M.H. Chung, Validity and reliability of the Indonesian version of the Pittsburgh Sleep Quality Index in adolescents. International Journal of Nursing Practice, 2021. 27(5): p. e12856. DOI: 10.1111/ijn.12856

18. Afandi, B., et al., The individualization of care for people with diabetes during Ramadan fasting: A narrative review. Ibnosina Journal of Medicine and Biomedical Sciences, 2020. 12(02): p. 98–107. DOI: 10.4103/ijmbs.ijmbs_49_20

19. Alfadhli, E.M., Higher rate of hyperglycemia than hypoglycemia during Ramadan fasting in patients with uncontrolled type 1 diabetes: insight from continuous glucose monitoring system. Saudi Pharmaceutical Journal, 2018. 26(7): p. 965–969. DOI: 10.1016/j.jsps.2018.05.006

20. Hosseini, S.S., et al., The effect of educational program based on theory of planned behavior on promoting retinopathy preventive behaviors in patients with type 2 diabetes: RCT. BMC Endocr Disord, 2021. 21(1): p. 17. DOI: 10.1186/s12902-021-00680-2

21. Briançon-Marjollet, A., et al., The impact of sleep disorders on glucose metabolism: endocrine and molecular mechanisms. Diabetology & metabolic syndrome, 2015. 7: p. 1–16. DOI: 10.1186/s13098-015-0018-3

